# Transmission Dynamics of Coronavirus Disease 2019 (COVID-19) in the World: The Roles of Intervention and Seasonality

**DOI:** 10.1101/2020.07.17.20156430

**Authors:** Shunxiang Huang, Lin Wu, Li Xu, Aihong Zhang, Li Sheng, Feng Liu, Long Zhou, Jing Li, Rongzhang Hao, Hua Qian, Sheng Fang, Zhongyi Wang, Yingru Li, Yuguo Li, Chan Lu, Qihong Deng

**Affiliations:** Institute of NBC Defense of PLA, Beijing 102205, China; State Key Laboratory of Atmospheric Boundary Layer Physics and Atmospheric Chemistry, Institute of Atmospheric Physics, Chinese Academy of Sciences, Beijing 100029, China; National Meteorological Center, China Meteorological Administration, Beijing 100081, China; Beijing Pure Blue Technology Co., Ltd., Beijing 100085, China; Aerospase information Research Institute, Chinese Academy of Sciences, Beijing 100094, China; School of Public Health, Capital Medical University, Beijing 100069, China; school of energy and environment, Southeast University, Nanjing 210096, China; Institute of Nuclear and New Energy Technology, Collaborative Innovation Centre of Advanced Nuclear Energy Technology, Key Laboratory of Advanced Reactor Engineering and Safety of Ministry of Education, Tsinghua University, Beijing, 100084, China; Institute of Biotechnology, Academy of Military Medical Sciences, Beijing 100071, China; Materials Institute, China Academy of Engineering Physics, Jiangyou, Sichuan 621908, China; Department of Mechanical Engineering, The University of Hong Kong, Hong Kong, China; XiangYa School of Public Health, Central South University, Changsha, Hunan 410078, China

**Keywords:** COVID-19, SARS-CoV-2, SEIR-CV, intervention effect, Asymptomatic infection, seasonal variation

## Abstract

The coronavirus disease 2019 (COVID-19) is spreading rapidly all over the world. The transmission dynamics of the COVID-19 pandemic is still unclear, but developing strategies for mitigating the severity of the pandemic is yet a top priority for global public health. In this study, we developed a novel compartmental model, SEIR-CV(susceptible-exposed-infectious-removed with control variables), which not only considers the key characteristics of asymptomatic infection and the effects of seasonal variations, but also incorporates different control measures for multiple transmission routes, so as to accurately predict and effectively control the spread of COVID-19. Based on SEIR-CV, we predicted the COVID-19 epidemic situation in China out of Hubei province and proposed corresponding control strategies. The results showed that the prediction results are highly consistent with the outbreak surveillance data, which proved that the proposed control strategies have achieved sound consequent in the actual epidemic control. Subsequently, we have conducted a rolling prediction for the United States, Brazil, India, five European countries (the United Kingdom, Italy, Spain, Germany, and France), southern hemisphere, northern hemisphere, and the world out of China. The results indicate that control measures and seasonal variations have a great impact on the progress of the COVID-19 pandemic. Our prediction results show that the COVID-19 pandemic is developing more rapidly due to the impact of the cold season in the southern hemisphere countries such as Brazil. While the development of the pandemic should have gradually weakened in the northern hemisphere countries with the arrival of the warm season, instead of still developing rapidly due to the relative loose control measures such as the United States and India. Furthermore, the prediction results illustrate that if keeping the current control measures in the main COVID-19 epidemic countries, the pandemic will not be contained and the situation may eventually turn to group immunization, which would lead to the extremely severe disaster of about 5 billion infections and 300 million deaths globally. However, if China’s super stringent control measures were implemented from 15 July, 15 August or 15 September 2020, the total infections would be contained about 15 million, 32 million or 370 million respectively, which indicates that the stringent and timely control measures is critical, and the best window period is before September for eventually overcoming COVID-19.

**Significance:** COVID-19 is now posing a huge threat to global public health. The key features such as asymptomatic infection and droplet or airborne transmission make COVID-19 more easily spread and more widely distributed around the world. It is an urgent need to explore the optimal intervention strategies and measures to contain the pandemic. Our novel SEIR-CV compartmental model considers the new features of COVID-19, exhibits the impact of the intervention strategies and seasonal variations, and thus can accurately predicts its trajectory in China and the rest of the world. Our research results suggest that control measures and seasonal variations have a great impact on the development of the COVID-19 pandemic, which can only be contained by stringent strategies during the best window period before September 2020 for eventually overcoming COVID-19, otherwise it would cause a severer global catastrophe.

## 1. INTRODUCTION

The coronavirus disease 2019 (COVID-19), caused by a novel virus of the β-coronavirus genus (SARS-CoV-2), has been spreading globally.^1,2^ As of 30 June 2020, it was reported by the World Health Organization (WHO) that there was a total of 10,185,374 cases and 503,862 deaths in the world. The WHO declared the outbreak of COVID-19 to be a pandemic on 11 March, escalating their previous announcement that the outbreak was a public health emergency of international concern (PHEIC) on 30 January. ^3^ It is the third coronavirus epidemic this century, and now COVID-19 is posing a huge threat and even a disaster to the global public health.^4,5^

Mounting evidence indicates that SARS-CoV-2 may originate from bats and can be transmitted from human to human.^6,7^ SARS-CoV-2 is one of the seven coronavirus species known to infect humans, including the recent Severe Acute Respiratory Syndrome coronavirus (SARS-CoV) and the Middle East Respiratory Syndrome coronavirus (MERS-CoV).^8^ However, the latest studies indicate that the infection of COVID-19 may be via multiple routes including contact, droplets and aerosol,^9^ especially a large number of asymptomatic infections with infectiousness; ^10-12^ these new features make it more contagious than SARS or MERS and more widely distributed around the world.^13^ In our previous studies we tested environmental samples in hospital wards in Wuhan, China, and found that the air and object surfaces in COVID-19 wards were widely contaminated by SARS-CoV-2, especially the viruses were detected in air ≈4 m from patients, which implied a potentially high infection risk for medical staff and other close contacts.^14^

Intervention strategies and seasonal variations are two basic factors that affect the development of the COVID-19 pandemic. ^15,16^ The COVID-19 pandemic is now spreading rapidly outside China and there is an urgent need to accurately predict its fate and trajectory. In previous study, compartmental mathematical models have been widely and successfully applied to predict epidemic spread of infectious diseases. As early as 1927, Kermack and Mckendrick developed the classic SIR model (Susceptible, Infectious, and Recovered) to predict the infectious diseases in which patients gain long term immunity after being cured.^17^ Then this model has widely used and extended in the modeling of various human infectious diseases. It is worth noting that Anderson et al. (1991) developed a SEIR (Susceptible, Exposed, Infectious, and Recovered) model to predict epidemics with incubation periods.^18^ The SEIR model and its updates were successful in predicting the course of the SARS epidemic in 2003-2004,^19^ the pandemic H1N1 influenza pandemic in 2009,^20^ and MERS epidemic in 2012-2015.^21^ Recent studies have tried to use the SEIR model to predict the spread of COVID-19 in China. Raed et al. (2020) found that there would be about 21,022 (11,090–33,490) total infections by February 22 in Wuhan,^22^ while Wu et al. (2020) estimated that there would be 75,815 COVID-19 cases (37,304-130,330) in Wuhan by 25 January 2020.^23^ However, these predicted results are much higher than the reported COVID-19 cases (less than 1,000) at the time. The huge discrepancy between the predicted and the reported data may be mainly due to two reasons: one is the new features of COVID-19 and the other is the impact of incorporated interventions. The above models usually assumed a single intervention strategy and constant control measures during the entire course of the epidemic,^24,25^ but Chinese government gradually adjusted the intervention strategies and the corresponding control measures according to the evolution of COVID-19. Mounting evidence has shown that the timing, duration and intensity of the intervention are key factors influencing the epidemic dynamics.^26^ However, incorporating the heterogeneous effects of different intervention strategies and measures in infectious disease models still remain a challenge.^27^It is still unclear how to quantify and distinguish the impact of interventions and seasonal variations on the epidemic situation^28^, and it is also unclear how much the intervention measures such as quarantine, isolation, protection, and decontamination have contributed to the prevention of the development of the COVID-19 pandemic. Besides, the new features of COVID-19 such as asymptomatic infection have not been considered in the above models.

To fill the gap of knowledge, we developed a novel SEIR model with control variables, SEIR-CV (Figure 1). We consider the key characteristics of asymptomatic infection and incorporate several control measures for the multiple transmission routes of COVID-19, including personal protection, decontamination for the potentially SARS-CoV-2 contaminating locations, quarantine for close contacts and the susceptible in outbreak areas at home or designated places, and isolation of infections and suspected infections in hospitals. (Figure1A). We then evaluated the impact of intervention strategies on the transmission of COVID-19 (Figure 2 and Table 1). By taking above advantages of the model, we accurately predicted the trajectory of COVID-19 in China, the United States, Brazil, India, five European countries (UK, Italy, Spain, Germany, and France), southern hemisphere, northern hemisphere, and the world out of China, and showed the key role of intervention and seasonal variations in the evolution of the pandemic. The results show that China has essentially succeeded in controlling COVID-19 by gradually implementing more and more stringent interventions. However, we predicted that the pandemic could only be contained only when precisely and adaptive stringent interventions will be implemented as soon as possible. Otherwise, it would result in a severer global catastrophe.

**Table 1.** Control variables for intervention strategies in SEIR-CV model.

| Intervention strategy | Quarantine $\lambda_Q$ | | Isolation $\lambda_I$ | | decontamination $\lambda_D$ | | Protection $\lambda_P$ | |
| --- | --- | --- | --- | --- | --- | --- | --- | --- |
|  | Target | Rate | Target | Rate | Target | Rate | Target | Rate |
| **China outside Hubei province** |  |  |  |  |  |  |  |  |
| Strategy I (21 January 2020)<br>*Wuhan's lockdown, Emergency level I response* | 0.813 | 0.864 | 0.957 | 0.883 | 0.106 | 0.583 | 0.712 | 0.810 |
| Strategy II (04 February 2020)<br>*Comprehensive closed community* | 0.863 | 0.975 | 0.990 | 0.954 | 0.198 | 0.783 | 0.823 | 0.936 |
| Strategy III (20 March 2020)<br>*Precise control gradually* | 0.672 | 0.832 | 0.983 | 0.961 | 0.101 | 0.534 | 0.460 | 0.531 |
| **The United States** |  |  |  |  |  |  |  |  |
| Strategy I (01 March 2020) | 0.595 | 0.950 | 0.839 | 0.815 | 0.041 | 0.652 | 0.133 | 0.895 |
| Strategy II (20 March 2020) | 0.682 | 0.923 | 0.878 | 0.925 | 0.112 | 0.679 | 0.442 | 0.895 |
| Strategy III (30 March 2020) | 0.721 | 0.912 | 0.904 | 0.927 | 0.124 | 0.786 | 0.623 | 0.887 |
| Strategy IV (09 April 2020) | 0.725 | 0.932 | 0.911 | 0.925 | 0.129 | 0.794 | 0.630 | 0.898 |
| Strategy V (19 April 2020) | 0.702 | 0.897 | 0.910 | 0.922 | 0.113 | 0.813 | 0.613 | 0.873 |
| Strategy VI (29 April 2020) | 0.681 | 0.824 | 0.906 | 0.892 | 0.102 | 0.856 | 0.602 | 0.893 |
| Strategy VII (09 May 2020) | 0.653 | 0.849 | 0.902 | 0.815 | 0.092 | 0.891 | 0.598 | 0.812 |
| Strategy VIII (19 May 2020) | 0.518 | 0.814 | 0.899 | 0.781 | 0.077 | 0.775 | 0.442 | 0.816 |
| Strategy IX (29 May 2020) | 0.595 | 0.831 | 0.923 | 0.862 | 0.116 | 0.817 | 0.479 | 0.786 |
| Strategy X (08 June 2020) | 0.421 | 0.783 | 0.882 | 0.789 | 0.078 | 0.853 | 0.325 | 0.821 |
| **Brazil** |  |  |  |  |  |  |  |  |
| Strategy I (01 March 2020) | 0.388 | 0.608 | 0.794 | 0.765 | 0.095 | 0.518 | 0.320 | 0.669 |
| Strategy II (22 March 2020) | 0.619 | 0.623 | 0.885 | 0.837 | 0.124 | 0.624 | 0.655 | 0.697 |
| Strategy III (19 April 2020) | 0.653 | 0.616 | 0.891 | 0.842 | 0.136 | 0.720 | 0.711 | 0.748 |
| Strategy IV (09 May 2020) | 0.685 | 0.659 | 0.910 | 0.836 | 0.137 | 0.719 | 0.720 | 0.815 |
| Strategy V (29 May 2020) | 0.721 | 0.783 | 0.924 | 0.827 | 0.139 | 0.796 | 0.754 | 0.878 |
| **India** |  |  |  |  |  |  |  |  |
| Strategy I (01 March 2020) | 0.611 | 0.954 | 0.912 | 0.659 | 0.107 | 0.506 | 0.430 | 0.698 |
| Strategy II (25 March 2020) | 0.579 | 0.916 | 0.907 | 0.713 | 0.099 | 0.572 | 0.429 | 0.738 |
| Strategy III (09 April 2020) | 0.685 | 0.827 | 0.921 | 0.789 | 0.123 | 0.658 | 0.548 | 0.629 |
| Strategy IV (24 April 2020) | 0.602 | 0.682 | 0.882 | 0.781 | 0.089 | 0.712 | 0.484 | 0.786 |
| Strategy V (04 April 2020) | 0.615 | 0.791 | 0.897 | 0.813 | 0.113 | 0.772 | 0.500 | 0.815 |
| Strategy VI (19 April 2020) | 0.567 | 0.791 | 0.879 | 0.813 | 0.103 | 0.772 | 0.487 | 0.815 |
| **Five Europe countries (The United Kingdom, Spain, Italy, Germany and France)** |  |  |  |  |  |  |  |  |
| Strategy I (21 February 2020) | 0.723 | 0.845 | 0.843 | 0.889 | 0.103 | 0.527 | 0.420 | 0.727 |
| Strategy II (06 March 2020) | 0.673 | 0.781 | 0.832 | 0.831 | 0.094 | 0.682 | 0.413 | 0.793 |
| Strategy III (16 March 2020) | 0.802 | 0.831 | 0.856 | 0.912 | 0.121 | 0.637 | 0.511 | 0.712 |
| Strategy IV (26 March 2020) | 0.821 | 0.891 | 0.891 | 0.897 | 0.135 | 0.618 | 0.573 | 0.753 |
| Strategy V (05 April 2020) | 0.856 | 0.889 | 0.911 | 0.897 | 0.139 | 0.718 | 0.691 | 0.789 |
| Strategy VI (15 April 2020) | 0.737 | 0.892 | 0.909 | 0.890 | 0.113 | 0.818 | 0.757 | 0.850 |
| Strategy VII (25 April 2020) | 0.801 | 0.781 | 0.922 | 0.827 | 0.127 | 0.892 | 0.652 | 0.781 |
| Strategy VIII (05 May 2020) | 0.736 | 0.879 | 0.918 | 0.878 | 0.116 | 0.832 | 0.594 | 0.816 |
| Strategy IX (15 May 2020) | 0.689 | 0.819 | 0.913 | 0.921 | 0.101 | 0.781 | 0.559 | 0.781 |
| Strategy X (25 May 2020) | 0.552 | 0.783 | 0.911 | 0.782 | 0.086 | 0.892 | 0.478 | 0.792 |
| **Northern Hemisphere** |  |  |  |  |  |  |  |  |
| Strategy I (21 January 2020) | 0.765 | 0.803 | 0.895 | 0.802 | 0.121 | 0.372 | 0.689 | 0.663 |
| Strategy II (14 February 2020) | 0.568 | 0.852 | 0.906 | 0.829 | 0.068 | 0.721 | 0.399 | 0.737 |
| Strategy III (24 February 2020) | 0.649 | 0.882 | 0.918 | 0.917 | 0.113 | 0.883 | 0.420 | 0.816 |
| Strategy IV (25 March 2020) | 0.719 | 0.715 | 0.936 | 0.763 | 0.125 | 0.783 | 0.549 | 0.637 |
| Strategy V (09 April 2020) | 0.720 | 0.716 | 0.938 | 0.831 | 0.126 | 0.791 | 0.569 | 0.729 |
| Strategy VI (29 April 2020) | 0.689 | 0.816 | 0.920 | 0.783 | 0.088 | 0.814 | 0.492 | 0.785 |
| Strategy VII (19 May 2020) | 0.635 | 0.872 | 0.912 | 0.725 | 0.075 | 0.894 | 0.455 | 0.873 |
| Strategy VIII (08 June 2020) | 0.535 | 0.832 | 0.908 | 0.798 | 0.071 | 0.812 | 0.391 | 0.837 |
| **Southern Hemisphere** |  |  |  |  |  |  |  |  |
| Strategy I (01 March 2020) | 0.461 | 0.648 | 0.656 | 0.758 | 0.095 | 0.372 | 0.261 | 0.546 |
| Strategy II (20 March 2020) | 0.732 | 0.868 | 0.879 | 0.945 | 0.153 | 0.835 | 0.619 | 0.831 |
| Strategy III (04 April 2020) | 0.721 | 0.863 | 0.884 | 0.884 | 0.119 | 0.857 | 0.621 | 0.817 |
| Strategy IV (29 April 2020) | 0.761 | 0.823 | 0.934 | 0.783 | 0.131 | 0.789 | 0.687 | 0.793 |
| Strategy V (09 May 2020) | 0.725 | 0.819 | 0.925 | 0.881 | 0.135 | 0.816 | 0.628 | 0.814 |
| Strategy VI (24 May 2020) | 0.768 | 0.783 | 0.932 | 0.786 | 0.147 | 0.862 | 0.711 | 0.846 |

**Figure 1.**
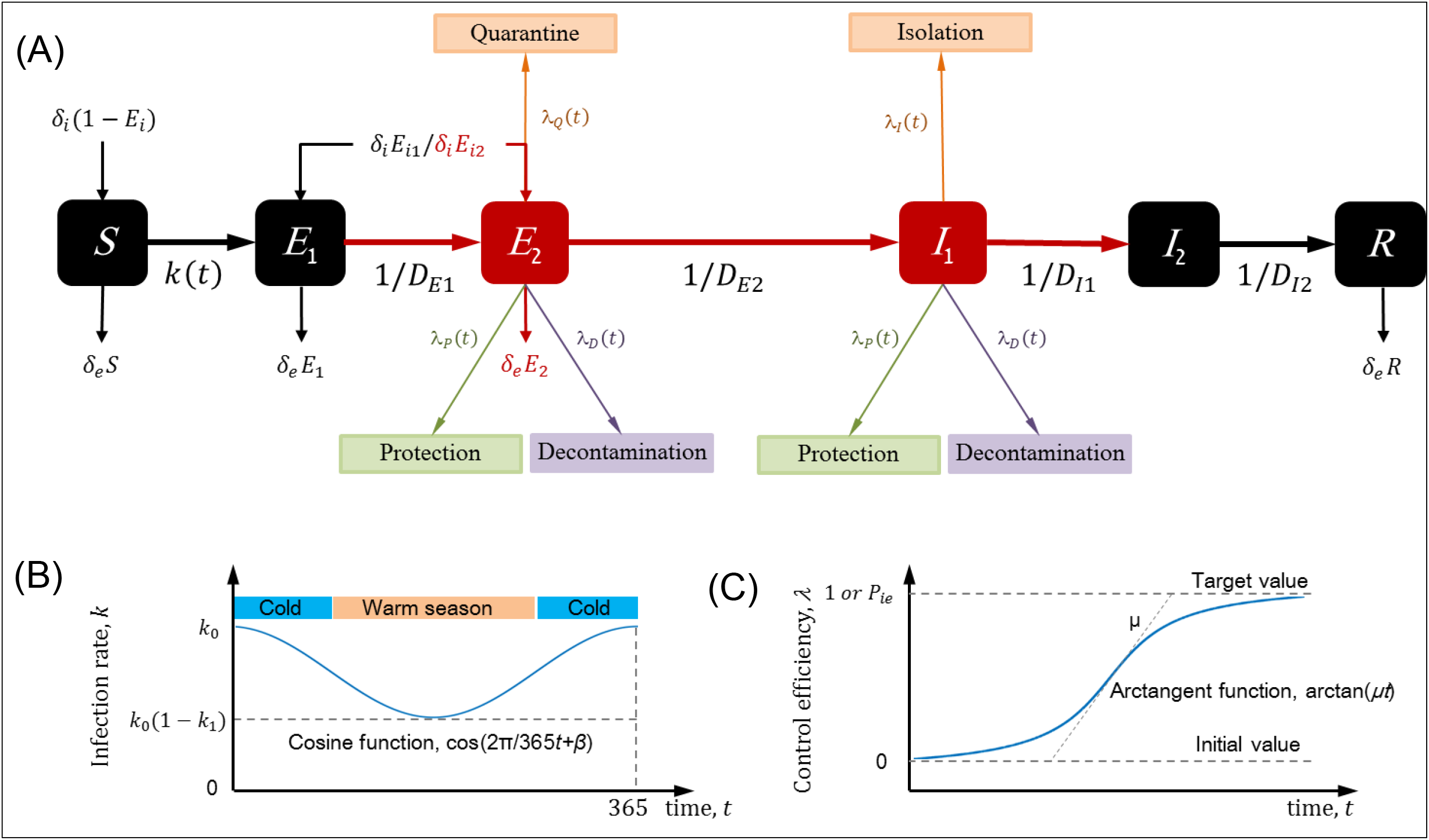
Compartmental model and intervention strategies. **(A) SEIR-CV compartmental model**. *S, E*_1_, *E*_2_, *I*_1_, *I*_2_ and *R* are respectively the fraction of susceptible, incubation with non-infectiousness, incubation with infectiousness, infection with infectiousness and infection with non-infectiousness, and recovered individuals, with *S*+*E*_1_+*E*_2_+*I*_1_+ *I*_2_+ *R* =1. *δ*_*i*_ is the immigration, *δ*_*e*_ is emigration rate, *E*_*i*_ is the proportion of asymptomatic infection from immigration, *E*_*i*1_ and *E*_*i*2_ is the proportion of non-infectious and infectious asymptomatic infections respectively, with *E*_i1_+*E*_*i*2_= *E*_*i*_; *k* is the infection rate, *k*_0_ is the basic infection rate, k1 is the seasonal coefficient; *D*_*E*1_, *D*_*E*2_, *D*_*I*1_ and *D*_*I*2_ are respectively durations of incubation with non-infectiousness, incubation with infectiousness, infection with infectiousness and infection with non-infectiousness. *λ*_*Q*_ is quarantine rate for the incubation with infectiousness, *λ*_*I*_ is isolation rate for the infect with infectiousness, *λ*_*P*_ and *λ*_*D*_ are respectively the decline of the infection rate due to prevention and decontamination measures, the constant β is 0 and π in the northern and southern hemispheres, respectively, *μ* is the implementation rate, and *P*_*ie*_ is the proportion of infections through the environment; **(B) The variation of infection rate with season**, *k*; **(C) The variations of the efficiencies of the control measures**, *λ*.

**Figure 2.**
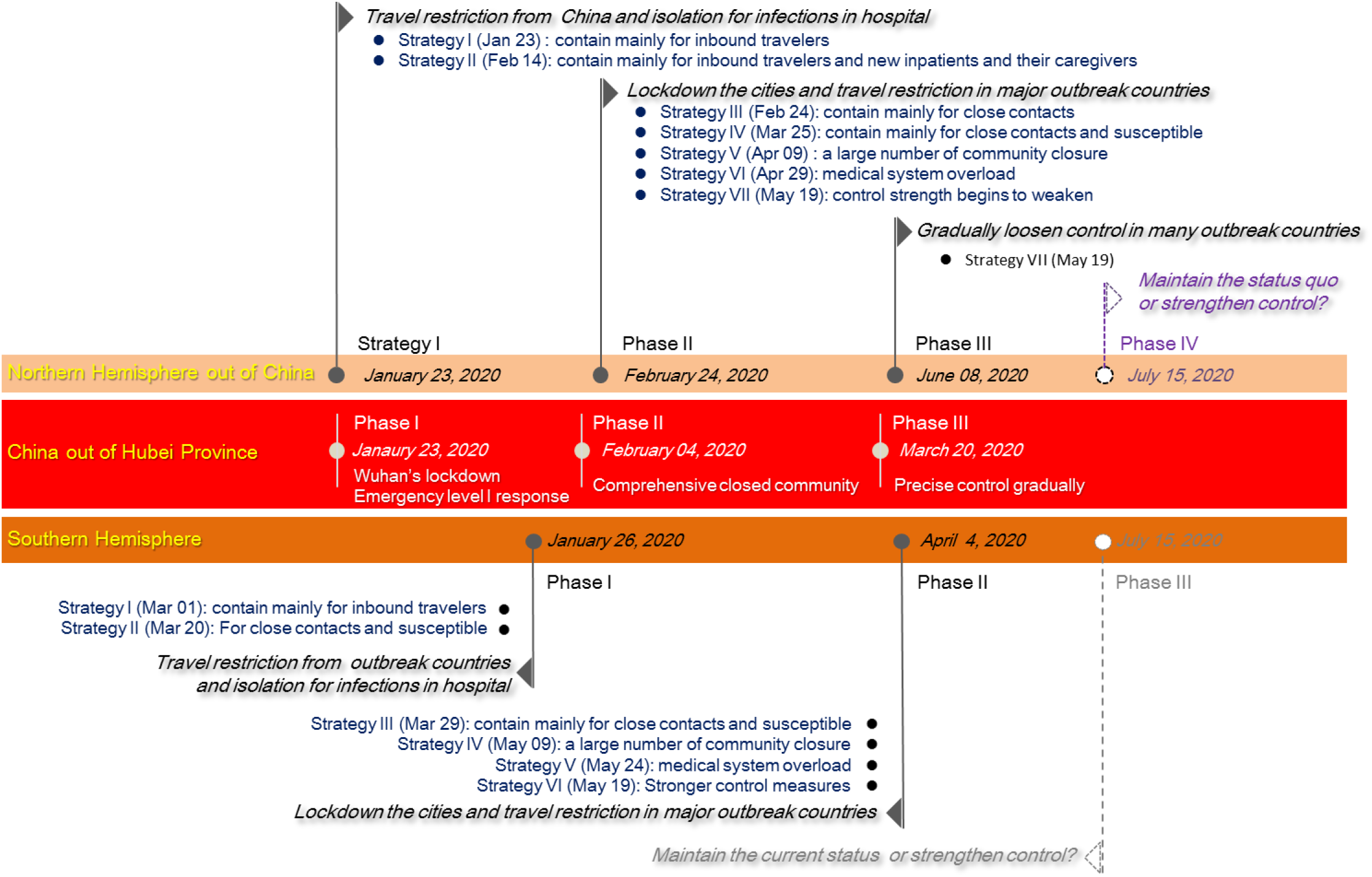
Intervention strategies implemented to contain COVID-19 in China out of Hubei province, southern hemisphere, and northern of out of China.

## 2. RESULTS

### 2.1 The Basic Infection Rate and Seasonal Influence

We inverted the parameters of the SEIR-CV for COVID-19 outbreak by the adjoint method in China out of Hubei, the United States, Brazil, five Europe countries (the United Kingdom, Spain, Italy, Germany and France), Northern Hemisphere, Southern Hemisphere, and the world out of China. The results show that the basic infection rate, *k*_0_, is 2.461∼2.650, which reveals the high infectivity of COVID-19. The regional differences are not obvious, though there is a relatively larger basic infection rate in the United States. The results also indicate that the seasonal coefficient of infection rate, *k*_1_, is 0.530∼0.698, and reflects that the infectivity of COVID-19 is greatly affected by seasonal variations, which have obvious regional differences, for example the Indian COVID-19 epidemic is less affected by seasonal variations(Table 2). Comparison results of the effects of seasonal variations and control measures on the COVID-19 outbreak in the United States and in Brazil indicate that control measures and seasonal variations have a great impact on the development of the epidemic. Assuming that there are no control measures or no seasonal variations in the United States, the inflection points with approximately 37 million and 15 million new cases may appear on 30 March 2020 and 30 May 2020, respectively, and group immunization may be formed in May and August. If there are no control measures in Brazil, the inflection points with approximately 23 million new cases may appear on 04 April 2020 and group immunization formed in May, but assuming that there are no seasonal variations in Brazil, the inflection points with only 208 new cases may appear on 04 April 2020 and the outbreak will end with 11359 total infections in the end of July on the contrary. (Figure 3 and the data table in Figure 3).

**Table 2.** Basic infection rate and Seasonal coefficient.

| Country | China out of Hubei | The United States | Brazil | India | Five European countries | Northern Hemisphere | Southern Hemisphere |
| --- | --- | --- | --- | --- | --- | --- | --- |
| Basic infection rate, $k_0$ | 2.509 | 2.650 | 2.596 | 2.466 | 2.530 | 2.461 | 2.533 |
| Seasonal coefficient, $k_1$ | 0.671 | 0.698 | 0.586 | 0.530 | 0.680 | 0.588 | 0.602 |

**Figure 3.**
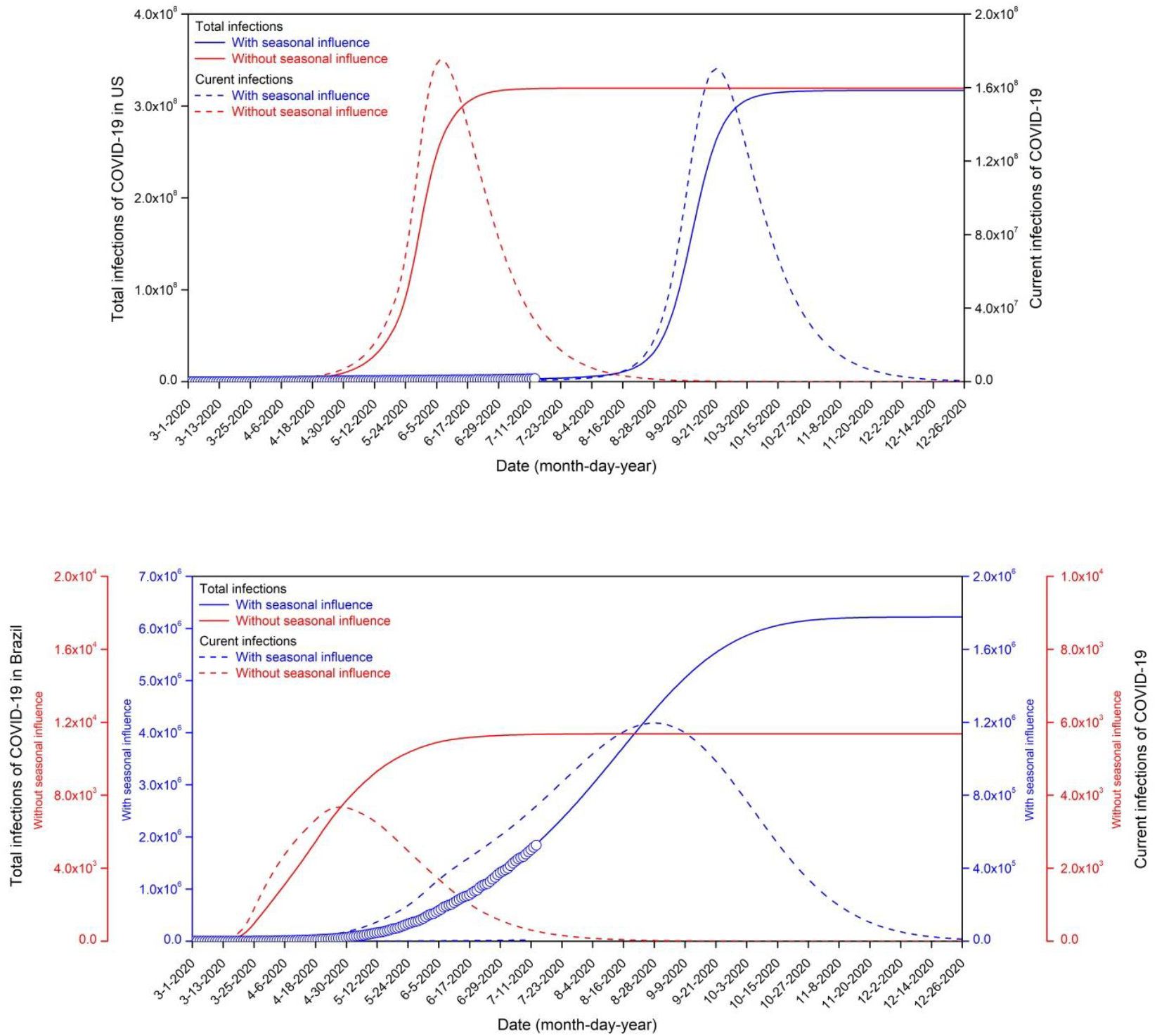
Comparison between the effects of seasonal variations and control measures on the COVID-19 outbreak in the United States and Brazil.

### 2.2 Comparison between rolling prediction and surveillance results

The rolling predicting are conducted on the trajectory of COVID-19 outbreak in China out of Hubei, the United States, India, Brazil, five Europe countries (the United Kingdom, Spain, Italy, Germany and France), Northern Hemisphere out of China, Southern Hemisphere, and the world out of China from January 21 to June 30, 2020 based on the outbreak surveillance results of the National Health Commission of the People’s Republic of China and WHO (Figure 4 and the data table on rolling prediction results), and the control variables through inversion update time is 10 to 40 days (Table S3) through inversion based on the outbreak surveillance results of the National Health Commission of the People’s Republic of China and WHO. Correlation analysis of rolling prediction and surveillance results show that the correlation coefficient, bias and average error are 0.99987∼0.99997, 9.7E-9∼5.0E-6 and 2.0E-6∼4.6E-5 for the total confirmed cases and the total prediction infected cases, 0.89934∼0.99658, 3.0E-6∼1.5E-3, 7.7E-4∼2.7E-2 for the total new confirmed cases and the total new prediction infected cases, respectively. And 95% confidence interval is 0.99976∼0.99999 for the total confirmed cases and the total prediction infected cases, 0.83728∼0.99793 for the total new confirmed cases and the total new prediction infected cases, respectively. (Table S2) The high consistency of all surveillance and prediction results reflects for different regions validates the accuracy and reliability of the SEIR-CV prediction.

**Figure 4.**
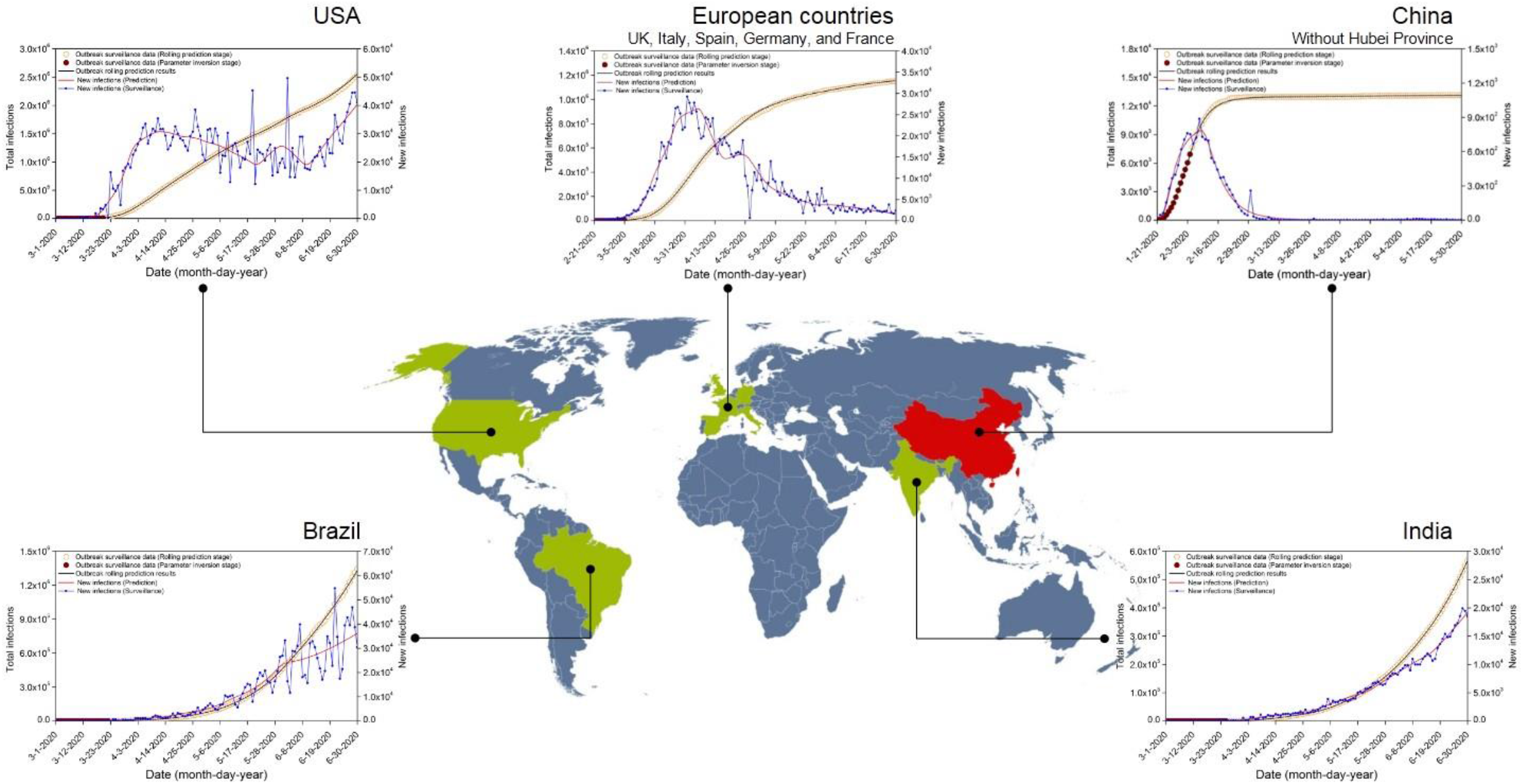
Comparison of rolling prediction and surveillance results for the COVID-19.

### 2.3 Pandemic period prediction for the COVID-19

We predicted the pandemic period of the COVID-19 in the world out of China, the United States, India and five European countries respectively.

#### (1) The COVID-19 outbreak in pandemic period in the United States

If the current intervention measures are maintained, the inflection point would appear on 12 September 2020 resulting in over 13 million new cases. The total infections would be up to 0.3 billion in the United States in early October 2020, and the COVID-19 pandemic would continue beyond the end March 2021. If the stringent control measures with the Strategy IV (9∼18 April 2020) (Table 1) are taken from 15 July 2020, the pandemic would weaken gradually to the lowest point on 14 September 2020 with 3840 new cases, and then the outbreak will again quickly develop along with the infection rate rising in the cold season, the inflection point would be delayed to 12 January 2021 with over 6.6 million new cases, resulting in over 0.26 billion total infections in the early March 2021. If the stringent control measures with the Strategy IV (9∼18 April 2020) are taken from 15 August 2020, the inflection point would be delayed to 10 December 2020 with over 3.9 million new cases, resulting in approximately 0.24 billion total infections in the end of January 2021. If the stringent control measures with the Strategy IV (9∼18 April 2020) are taken from 15 September 2020, there would be over 0.28 billion total infections in the early October 2020. If the China’s super-stringent control measures are taken in the United States from 15 July 2020, 15 August 2020 and 15 September 2020, respectively, there would be approximately 3.5 million total infections in the middle August 2020, 15 million total infections in the middle November 2020 and 260 million total infections in the middle November 2020, respectively. (Figure 5 and the data table in Figure 5)

**Figure 5.**
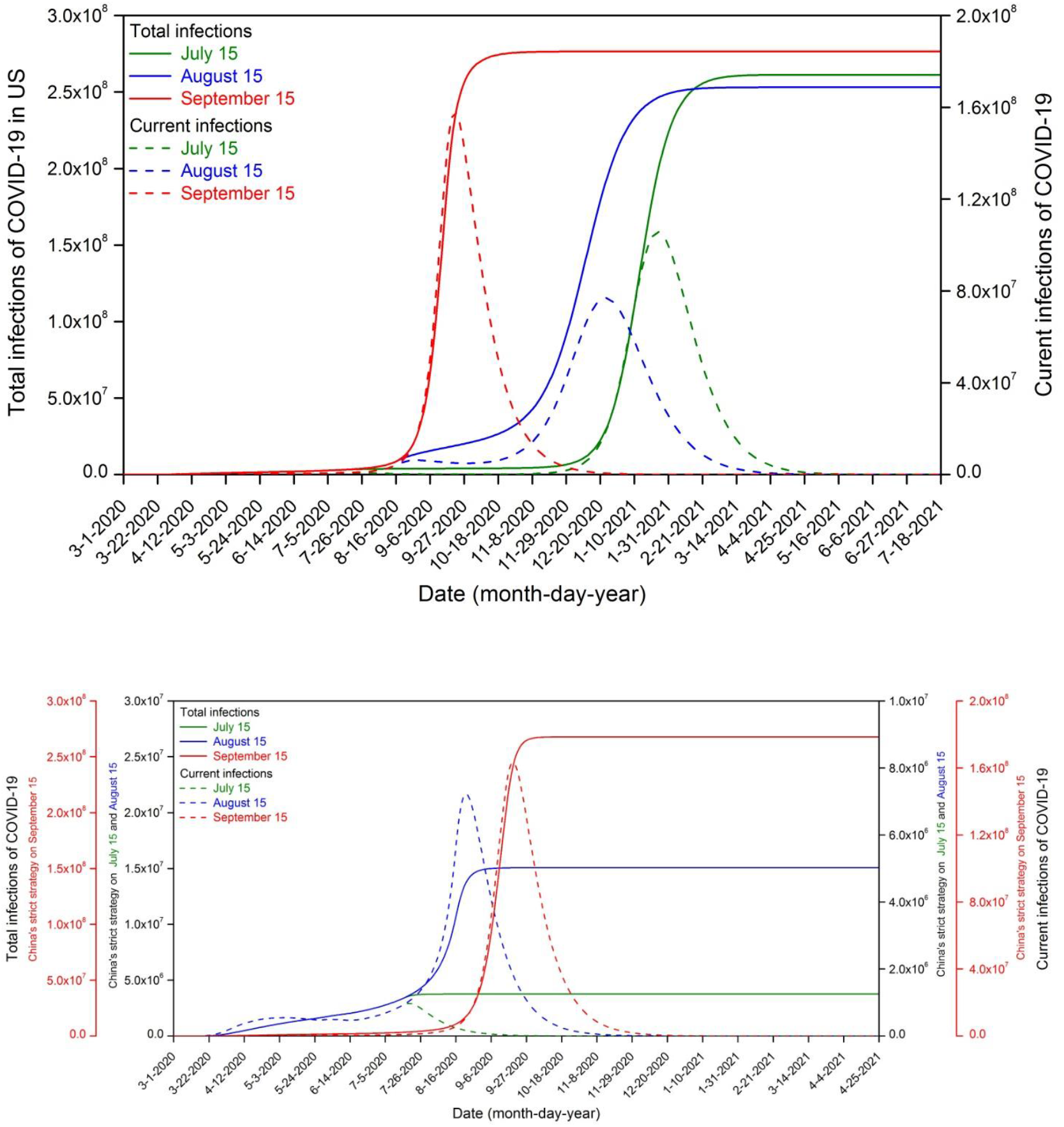
Prediction of the COVID-19 in the United States. (A) Implement current control measures. (B) Implement strict control measures already taken in the United States from 15 July 2020. (C) Implement strict control measures already taken in the United States from 15 August 2020. (D)Implement strict control measures already taken in the United States from 15 September 2020. (E) Implement China’s super-strength measures from 15 July 2020. (F) Implement China’s super-strength measures from 15 August 2020. (G) Implement China’s super-strength measures from 15 September 2020.

#### (2) The COVID-19 outbreak in pandemic period in India

If the current intervention measures are maintained, the inflection point would appear on 10 October 2020 resulting in over 47 million new cases. The COVID-19 pandemic would continue beyond the end March 2021, and the total infections will be up to 1.24 billion in India in the middle December 2020. If the stringent control measures with the Strategy IV (9∼23 April 2020) (Table 1) are taken from 15 July 2020, the pandemic would be weaken gradually to the lowest point on 24 August 2020 with 12770 new cases, and then the outbreak will again quickly develop along with the infection rate rising in the cold season, the inflection point would be delayed to 23 December 2020 with over 30 new million infections, resulting in over 1 billion total infections in the middle January 2021. If the stringent control measures with the Strategy IV (9∼23 April 2020) are taken from 15 August 2020, the inflection point would appear on 04 December 2020 with over 27 million new cases, resulting in over 1 billion total infections in the end of December 2020. If the stringent control measures with the Strategy IV (9∼23 April 2020) are taken from 15 September 2020, the inflection point would appear on 4 November 2020 with over 15 million new cases, resulting in over 0.9 billion total infections in the early December 2020. If the stringent control measures with the Strategy IV (9∼23 April 2020) are taken from 15 October 2020, the total infections will be resulted in over 1.1 billion in the end of October 2020. If the China’s super-stringent control measures are taken in India from 15 July 2020, 15 August 2020 and 15 September 2020, respectively, there would be approximately 1.2 million total infections in the early September 2020, 4.4 million total infections in the middle October 2020 and 74 million total infections in the end of December 2020, respectively.(Figure 6 and the data table in Figure 6)

**Figure 6.**
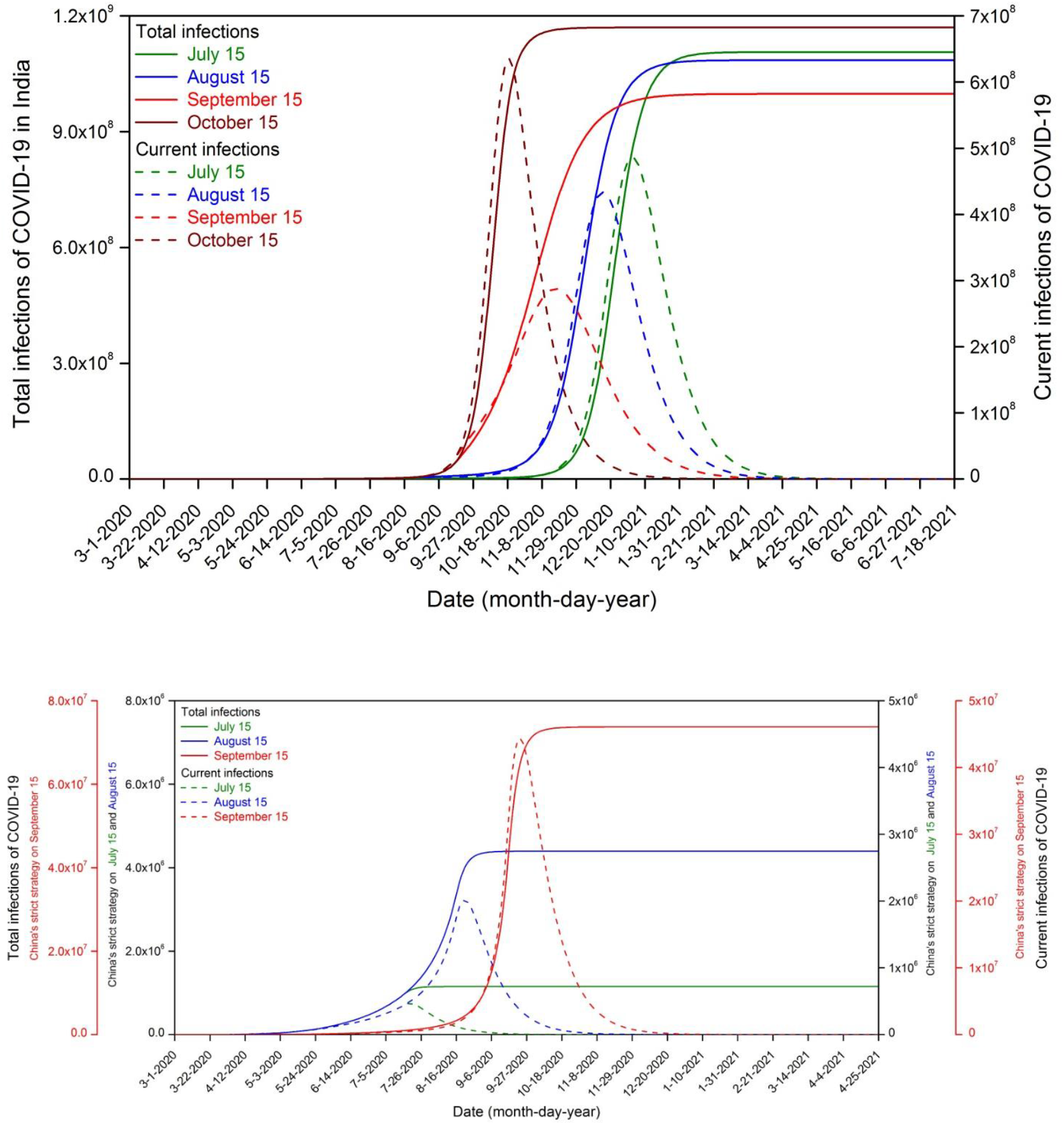
Prediction of the COVID-19 in India. (A) Implement current control measures. (B) Implement strict control measures already taken in India from 15 July 2020. (C) Implement strict control measures already taken in India from 15 August 2020. (D)Implement strict control measures already taken in India from 15 September 2020. (E) Implement China’s super-strength measures from 15 July 2020. (F) Implement China’s super-strength measures from 15 August 2020. (G) Implement China’s super-strength measures from 15 September 2020.

#### (3) The COVID-19 outbreak in pandemic period in five European countries

If the current intervention measures are maintained, the inflection point would appear on 7 November 2020 resulting in about 15 million infections. The COVID-19 pandemic would continue beyond the end March 2021, and the total infections will be up to 0.3 billion in five European countries in early December 2020. If the stringent control measures with the Strategy V (5∼14 April 2020) (Table 1) are taken from 15 July, 2020, the pandemic would weaken gradually to the end in early June, 2021 with about 1.2 million infections. If the stringent control measures with the Strategy V (5∼14 April 2020) are taken from 15 August and September, 2020, respectively, the pandemic would be weaken gradually to the lowest point on November 02 with 40 infections and November 05, 2020 with 2282 infections, and then the outbreak will again quickly develop along with the infection rate rising in the cold season, the inflection point would be delayed to February 17 with 147 infections and 16 February, 2021 with 8410 infections, resulting in 1.2 million and 2.3 total infections in the end of April 2021, respectively. If the stringent control measures with the Strategy V (5∼14 April 2020) are taken from 15 October, 2020, the inflection point would appear on 4 January 2021 with over 0.5 million infections, resulting in over 70 million total infections in the end of February 2021. If the China’s super-stringent control measures are taken in five European countries from 15 July 2020, 15 August 2020, 15 September 2020 and 15 October respectively, the outbreak will end with approximately 1.19 million total infections in the early September 2020, 1.20 million total infections in the early October 2020, 1.33 million total infections in the early December 2020, and 10.7 million total infections in the middle February 2021, respectively. (Figure 7 and the data table in Figure 7)

**Figure 7.**
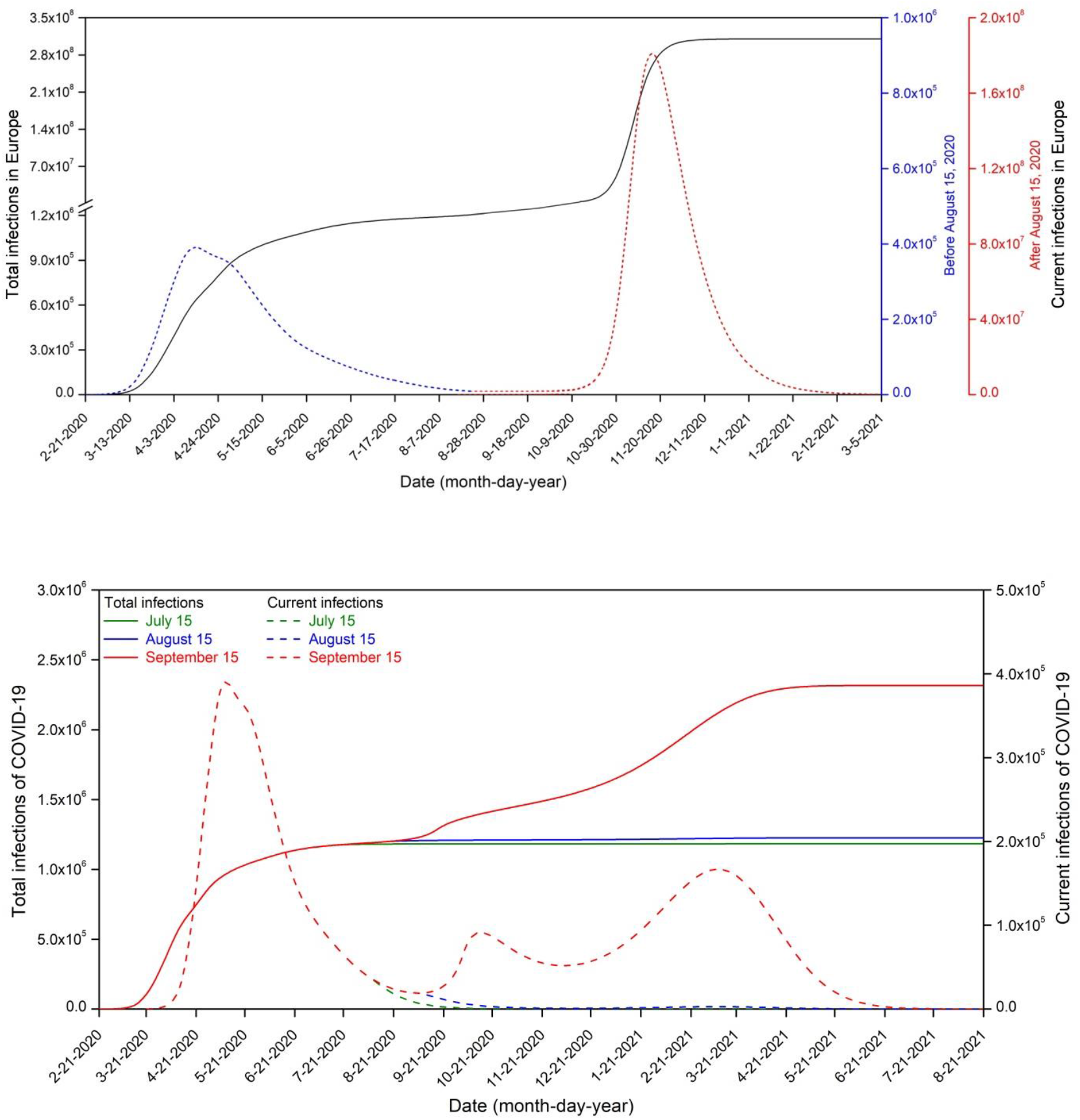
Prediction of the COVID-19 in five European countries. (A) Implement current control measures. (B) Implement strict control measures already taken in five European countries from 15 July 2020. (C) Implement strict control measures already taken in five European countries from 15 August 2020. (D)Implement strict control measures already taken in five European countries from 15 September 2020. (E) Implement China’s super-strength measures from 15 July 2020.

#### (4) The COVID-19 outbreak in pandemic period in the world out of China

If the current intervention measures are maintained, the inflection point would appear on 7 October 2020 resulting in over 205 million new cases, the peak of latent persons and infections would occur in the early October and in the middle October 2020 resulting in 1.0 billion and 2.7 billion cases, respectively. The COVID-19 pandemic would continue beyond the end April 2021, and the total infections would be up to 5 billion in the world out of China in early November 2020. If the stringent control measures with the Strategy V (9∼28 April 2020) (Table 1) in northern hemisphere out of China and the Strategy VI (24 May ∼ 29 June 2020) (Table 1) in southern hemisphere are taken from 15 July 2020, the pandemic would weaken gradually to the lowest point on 17 October 2020 with 47700 new cases, and then the outbreak would again quickly develop along with the infection rate rising in the cold season. In this case, the inflection point would be delayed to 5 February 2021 with over 8 million infections, resulting in approximately 3.6 billion total infections in the end of April 2021. Assuming that the China’s super-stringent control measures are taken from 15 July 2020, 15 August 2020 and 15 September 2020 respectively, the pandemic will end with approximately 15 million total infections in the end of October 2020, 33 million total infections in the middle December 2020, and 370 million total infections in the middle February 2021, respectively. (Figure 8)

**Figure 8.**
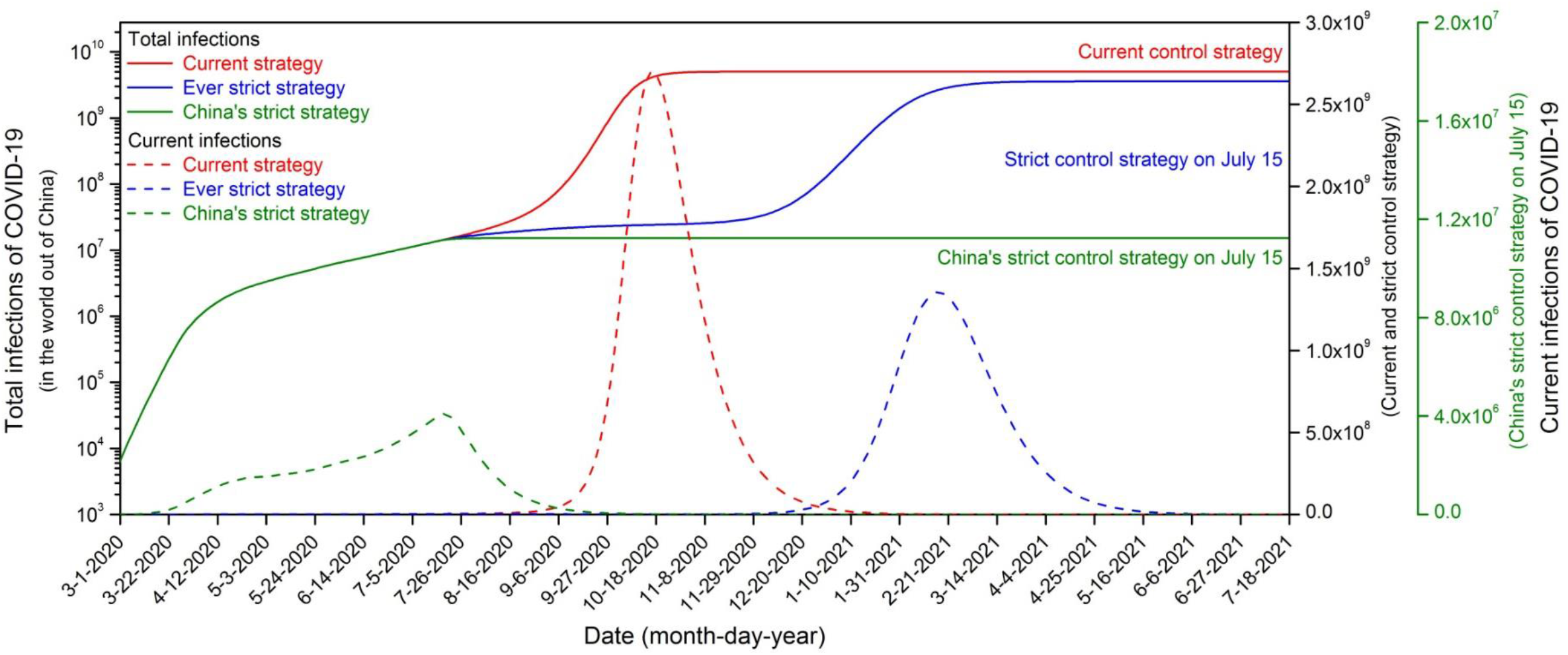
Prediction of the COVID-19 in the world out of China. (A) Implement current control measures. (B) Implement strict control measures already taken in five European countries from 15 July 2020. (C) Implement China’s super-strength measures from 15 July 2020.

## 3. DISCUSSION

We developed a novel SEIR-CV compartmental model, i.e,. SEIR-CV model, to predict the evolution of COVID-19 pandemic in both China and the rest of the world, and evaluated the impact of intervention strategies and seasonal variations on the disease transmission. Our model successfully predicted that the COVID-19 epidemic would be contained in China and would end in June except for the second outbreak for viruses SARS-CoV-2 from outside in the seafood market in Xinfadi, Beijing. The new model also predicted the rapid spreading of COVID-19 outside China. Our prediction results indicate that control measures and seasonal variations are the two key factors that affect the development of the COVID-19 pandemic. Strategies to treat all infections in emergency and/or shelter hospitals are urgently needed so that the COVID-19 epidemic can finally be contained.

In the northern hemisphere excluding China, the outbreak countries are currently in the warm season, which should have been the best window period for controlling the pandemic. However, the COVID-19 pandemic has not been contained because of the current weak intervention strategy. The COVID-19 infectivity will increase as the season becomes colder. Consequently, if the control measures are not strengthened, the pandemic will develop very rapidly and the total infections may be over 5 billion in the end of this year, which will continue beyond the end of April 2021. Of course, on the one hand, these countries’ medical systems will overload with the emergence of a large number of infections and many infections without effective treatment will worsen the epidemic situation. On the other hand, some countries with the rapid outbreak of the pandemic usually implement more stringent control measures to prevent the development of the outbreak and to reduce the total infections. In fact, once the pandemic deteriorates sharply, people will automatically quarantine at home or strengthen their protective measures for their own health and safety. Therefore, it is a small-probability event that the total infections quickly increase to 5 billion to acquire the group’s immunity.

In the southern hemisphere, COVID-19 is more contagious due to the cold season, so the current intervention strategies are still unable to effectively control the rapid development of the pandemic. Correspondingly, the current control intensity in the five European countries is weaker than that of Brazil, but it has been controlled to a certain extent because of the warm season reason. The infectivity will be weak with the temperature getting warmer. If the control measures are not weakened, the pandemic will gradually be controlled by the end of this year, and the total infections will reach over 10 million. However, if super-stringent control measures are taken like China, the epidemic situation will be quickly controlled; otherwise, if the control measures are relaxed later, it will bring more serious epidemic situation.

The novel SEIR-CV model presents its advantages that it explicitly incorporates the roles of different intervention measures and season. This enables us to determine the best strategy to control the dynamic evolution of COVID-19 by quantitatively involving the impact of seasonal variations on the epidemic situation, which can be transformed into achievable public health policies in time to mitigate the burden of the pandemic. (1) According to current measures, the United States cannot control the COVID-19 pandemic and will inevitably lead to a second huge outbreak. If China’s super-strong control measures are adopted before early September 2020 in the United States, it is still very effective to contain the COVID-19 pandemic. (2) According to current measures, it is very dangerous for India if the COVID-19 pandemic cannot be contained, which may cause 1.2 billion infections, and lead to the world’s worst pandemic. If China’s super-strong control measures are implemented before early October in India, it is still very effective to contain the COVID-19 pandemic. (3) According to the current measures of the five European countries, the epidemic will slowly develop in the near future, but it will rebound after entering the autumn and winter, and eventually towards group immunization. If China’s super-strong control measures are adopted, it is still very effective to contain the pandemic and August and September are the best window periods for epidemic control. (4) If China’s super-strong control measures are implemented in the world out of China, the COVID-19 pandemic can be contained and will end in this year with approximately 15.2 million total infections; If the current control measures are maintained, the COVID-19 pandemic will break out this fall and eventually approximately 5 billion infections; if stricter control measures (intermediate strategies) are implemented, the pandemic will be contained to a certain degree this year, but a second outbreak will be ushered in next spring, which may offer again important time and opportunities for the COVID-19 pandemic control.

Four types of control measures: quarantine, isolation, protection and decontamination were considered in our study. Case isolation and contact tracing is widely accepted protocols to contain the spread of infectious diseases,^29^ but is mainly useful for diseases transmitted by contact, such as the SARS outbreak in 2003 ^30^ and the Ebola outbreak in Africa in 2014.^31^ However, in addition to contact, COVID-19 can also be transmitted by respiratory droplet and even by air or airborne. Jiang et al. detected live SARS-CoV-2 viruses on surfaces and in the air of an isolation ward, indicating that transmission may happen via this route too.^32^ In our previous study, we found that the air and object surfaces in COVID-19 wards were widely contaminated by SARS-CoV-2.^14^ According to our epidemiological survey results, the ratio of cases from environment-to-person is 0.2. On the other hand, the COVID-19 infection may begin before the onset of symptoms.^7,11-12,33-35^ Case isolation alone is not sufficient to control COVID-19.^25,36^ There is an urgent need to quarantine exposed individuals without symptoms (self-isolation, home confinement, or community closure) and to take infection control measures (protection and decontamination) to prevent droplet and air transmission. However, the roles of quarantine, protection and decontamination have been greatly underestimated. Even with the rapid spread of COVID-19, many countries have not encouraged the use of face masks.

Our model considered the role of asymptomatic individuals on COVID-19 transmission. Some studies of 17 COVID-19 patients found that peak viraemia was observed at the end of the incubation period, indicating the possibility of transmission before onset of symptoms.^11-12,37^ Asymptomatic infection has been observed in several diseases, including malaria, MERS-CoV and Influenza A.^38-40^ However, in this aspect COVID-19 is different from SARS which little or no pre-symptomatic infectiousness was detected,^41^ even though the whole SARS-CoV-2 genome shares about 86% of the SARS-CoV genome.^42^ The characteristics of asymptomatic infection of COVID-19 means that COVID-19 is much easier to transmit and will be more widely distributed around the world. In our research, we have confirmed cases as infected cases, but the actual diagnosis time is related to the monitoring method, and some may have not been diagnosed when there are symptoms, for example, in the early epidemic in Wuhan, China. Persons may also be confirmed during incubation period, for example Chinese government makes nucleic acid testing compulsory among all key groups and available for those who ask to be tested, and some asymptomatic infections are included in the confirmed cases. In fact, the different monitoring methods may affect the consistency of the prediction results and the surveillance data, but have no effect on the prediction of the epidemic development trend. The surveillance results were higher than the prediction results, which illustrates this point (Figure 4).

Our study shows that the dynamic evolution of COVID-19 pandemic is highly sensitive to the intensity and timing of interventions, and early implementation of stringent intervention measures can effectively reduce the scale and duration. This is consistent with several other studies. For example, Yang et al. (2020) predicted, using machine learning, that a five-day delay in implementation would have increased the size of the epidemic in mainland China by three-fold.^24^ We also found that if the intensity of the control measures is not enough, even though the same control intensity, it only affects the time of the rapid outbreak, but eventually toward group immunization. Moreover, the total infections may be larger, even if the control measures are taken early. This is because the proportion of infections for group immunization is (1-1/*R*_0_), while *R*_0_ varies with seasons, so the total infections will be relatively small when the group immunization forms just during the warm season. However, this is the first study to examine the effect of intervention strategies including quarantine, isolation, protection and decontamination on the evolution of COVID-19. We found that if more strict control measures are not implemented, the COVID-19 pandemic will not end this year, but will be about to outbreak at a faster rate.

In fact, there might be more total cases than the actual surveillance data such as in Wuhan, but the model prediction using the surveillance data is consistent with other modeling results: Reed et al. (2020) predicted 21,022 (11090–33490) total infections before February 22;^5^ Roosa et al. (2020) predicted a cumulative number of reported cases of between 37,415 and 38,028 in Hubei Province before February 9;^43^ and Wang et al. (2020) estimated the number of infections in Wuhan would peak in late February with 58,077–84,520 cases.^44^ The difference may be due to the fact that many infected cases were undetected when there were no or very mild symptoms because of the limitation of the hospital’s admission conditions and diagnosis reagents in the early stage. However, our study indicates that the lockdown of Wuhan city during late January 2020 played a significant role in curbing the spread of COVID-19 in China and even the world. On one hand, implementation of travel restrictions within the city and shutting down transportation out of the city led to a remarkable decline in the spread of COVID-19 to other regions of China and the rest of the world.^45-47^ On the other hand, the Wuhan restrictions provide valuable response time and experience for controlling the disease. We see an obvious time lag between the Wuhan and other regions of China, and the rest of the world.

Our modeling indicates that the COVID-19 pandemic in the world outside of China is rapidly spreading. Urgent steps need to be taken by countries, particularly those with weak health care systems.^48^ The situation is really dangerous as the recent Lancet editorial “COVID-19: too little, too late?” published on March 2020 argued, that “countries have done too little, too late to contain the epidemic”.^49^ Some countries are still taking loose control measures. According to a recent WHO-China joint mission report, China’s measures to control the COVID-19 epidemic are the most “ambitious, agile and aggressive disease containment effort in history”, avoiding a large number of cases. The successful experience is encouraging many countries to deal with COVID-19 aggressively. Our modeling shows that the COVID-19 pandemic can only be contained this summer if other countries are willing to follow China’s stringent intervention measures, including the recruitment of medical staff and the establishment of emergency and/or shelter hospitals. However, it is unclear whether other countries will be able to implement the stringent measures that China eventually adopted.^50^ The world is now at a crossroads, and the attitude or the choice of each country will determine the outcome of COVID-19. It is now exciting that many countries are considering to implement more strict measures.

However, limitations in this study shoudl also be discussed. First, our prediction for the dynamics of COVID-19 spread does not consider the roles of vaccines. Development of vaccine is now underway, but it will still take a long time to be approved for use.^51^ Second, our modeling considers the five European countries and the southern hemisphere as a whole, and all countries were assumed to be taking the same measures, but the heterogeneity between these countries and travel between them might affect the prediction results.^45,46^ Third, we do not consider social events or behaviors. In western countries, there are more social activities during spring but less activities or events during winter when most people are at home, which is helpful for containing COVID-19. Fourth, we predicted a much stronger second outbreak without considering the improved responses of the government and the people to the pandemic. However, many people will learn from this pandemic and will be more careful and then keep distance and avoid large gatherings and events. Fifth, the parameters of our model were traced using adjoint method based on the surveillance data, but are not based on physical measurements. However, it is now impossible to obtain the values of parameters, for example the decontamination rate, by experiments. Lastly, we use the COVID-19 confirmed cases by WHO Coronavirus disease(COVID-19) Situation Report. In fact, some asymptomatic infections are not included in these data because they are not confirmed. Therefore, the actual infections should be more at present, and the actual confirmed cases in the future will also be less than the actual infections, but under the corresponding control measures, the predicted infections will be close to the actual infections.

In summary, a novel compartmental model, SEIR-CV, was developed to predict the spread of COVID-19 in China and the rest of the world, and to examine effectiveness of different intervention strategies are for containing the pandemic. Our findings indicate that the control strategies that are updated in a timely and effective manner is critical to succeeded in controlling COVID-19, such as those great efforts in China. These results provide a good template for other countries in the world to follow, where COVID-19 is rapidly spreading. As Maria van Kerkhove, the technical lead for the Health Emergencies Program at the WHO, said at a daily briefing in Geneva, Switzerland on March 3 2020: “Having China share its experience with other countries is nothing short of excellence”.

## 4. METHODS

### 4.1 SEIR-CV compartmental model

According to the COVID-19 characteristics, we developed a new compartmental model,

SEIR-CV, based on the classic SEIR (Susceptible-Exposed-Infectious-Recovered) model (Figure 1), which considered the following:

- The characteristics of asymptomatic infectiousness of the incubated individuals; ^10-12^
- Different measures for controlling the multiple infection routes, including quarantine at home or within communities(maintain social distance), isolation for infected individuals in hospitals, decontamination (such as sterilization, inactivate, washing hands) for polluted environment by SARS-CoV-2, and personal protection (such as wearing mask or protective suit, increasing ventilation);
- The seasonal variation of the transmission.

The differential equations are:

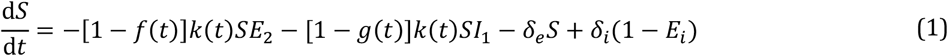

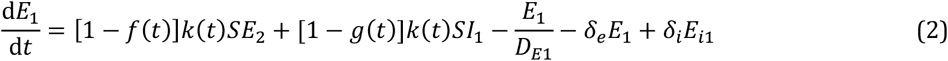

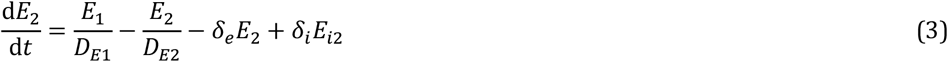

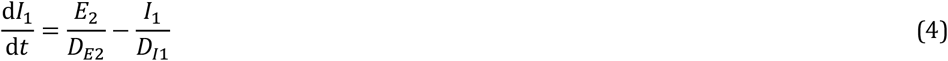

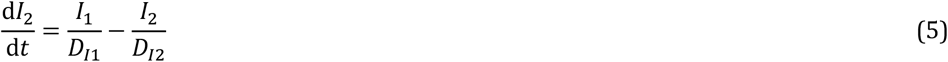

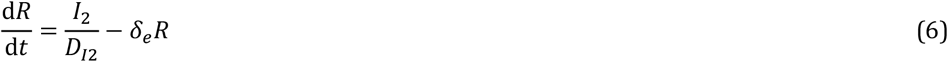

where, *δ*_*e*_ and *δ*_*i*_ are the proportion of emigration and immigration population, respectively; *E*_*i*_ is the proportion of asymptomatic infection from immigration, *E*_*i*1_ and *E*_*i*2_ is the proportion of non-infectious and infectious asymptomatic infections respectively. The infection rate, *k*, is considered to vary according to the seasons, higher during the cold season but lower during the warm season (Figure 1b) ^15,16,28^

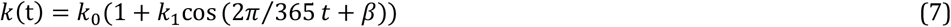

where *k*_0_ is the basic infection rate, related to factors such as the characteristics of the virus and the density of the population, assuming the same value of the asymptomatic and the symptomatic considering there being no obvious difference in viral loads across sex, age groups and disease severity^10^, *k*_1_ is the seasonal coefficient.

Contagious function, and *β*is 0 and *π*in the northern and southern hemispheres, respectively.

The effects of the control measures on the disease transmission are described by the following functions:

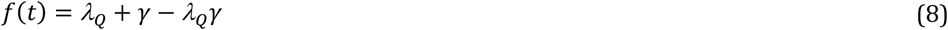

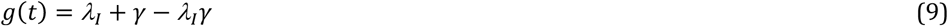

where *λ*_*Q*_, *λ*_*I*_, *λ*_*P*_, *λ*_*S*_ are the efficiencies of the control measures. They are independent of each other, described as follows, Figure 1c:

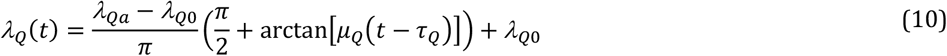

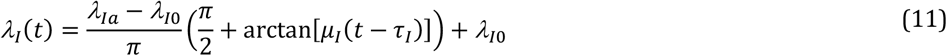

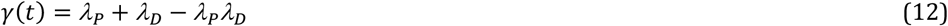

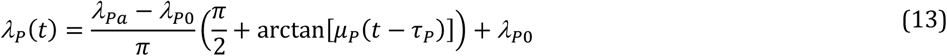

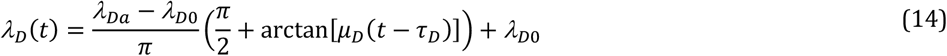

where *λ*_*Qa*_, *λ*_*Ia*_, *λ*_*Pa*_, *λ*_*Da*_ are respectively, the target efficiencies of quarantine, isolation, protection and decontamination, and *λ*_*Q*0_, *λ*_*I*0_, *λ*_*P*0_, *λ*_*D*0_ are the initial efficiencies; *μ* is the implementation rate and *τ* is the time of the intervention measures. Since the effect of an intervention is usually gradual, we assumed that the dynamic process conforms to an arctangent function, and thus the effect of intervention will be asymptotic to the target values: the higher the implementation rate (*μ*), the faster the target value can be reached (Please see Figure 1c).

### 4.2 Intervention strategies

To better control the spread of infectious disease, different intervention strategies should be implemented in response to the course of an epidemic. According to the evolution of COVID-19, governments have progressively adjusted their intervention.

*(1) Strategy I (January 23, 2020): Wuhan’s lockdown and Emergency level I response. The government decided to restrict travel in the city and shut down transportation out of the city. Control measures including isolation, protection and decontamination, were implemented nationwide. Since WHO declared COVID-19 to be a public health emergency of international concern, stringent prevention and control measures (major public health emergency level I response) were implemented in all provinces and cities throughout mainland China. Public places were closed and public services were banned and transportation across cities was halted*.

*(2) Strategy II (February 04, 2020)*: *Comprehensive closed community*. Self-isolation and home confinement management. As the infection continues and rapidly increases, all communities were closed and people were required to stay at home in order to reduce the possibility of contact with exposed or infected individuals.

*(3) Strategy III (March 20, 2020): Precise control gradually*. Chinese government began to allow the resumption of work progressively across the whole country with the exception of the major epidemic areas. However, to avoid an increase in the epidemic, all the communities implemented self-isolation management. Due to the declining trend of epidemic, the central government decided to implement scientific but precise prevention and control measures, and required all localities to adjust the emergency response level based on actual situation so as to restore normal production and life as early as possible.

### 4.3 Parameterization and solution

A main challenge of modeling the epidemics of infectious diseases is the difficulty in determining the parameters in the compartmental models. In the present study, we used an adjoint method to identify the parameters in the SEIR-CV model, because that its efficiency and robustness are critical for operational usage. (Please see the details in Supplemental Materials 2). Other more sophisticated methods such as Bayesian inference^52,53^ may also work with the SEIR-CV model and may provide additional statistical information in mathematics, but the increased computational cost might not be acceptable for operational usage.

The parameters of the SEIR-CV model are summarized in Table S4. According to the report about temporal patterns of viral shedding in 94 patients with laboratory-confirmed COVID-19, in which COVID-19 infectiousness profiles from a separate sample of 77 infector–infectee transmission pairs were modeled(He et al, 2020), non-infectious incubation period, *D*_E1_ is 2.9 days, infectious incubation period, *D*_E2_ is 2.3 days, infectious cure period, *D*_I1_ 7.0 day, and non-infectious cure period, *D*_I2_ is 12.0 days for COVID-19. The emigration/immigration rate, *δ*_*e*_/*δ*_*i*_, based on the data from the database of Baidu (http://qianxi.baidu.com/). From January to February 2020, we carried out the epidemiological survey in Wuhan, Hubei Province, China. A total of 20 confirmed patients were tracked and sorted out, including 4 cases from environment-to-person transmission and 16 cases from person-to-person transmission. According to the comprehensive epidemiological survey data, the ratio of cases from environment-to-person or person-to-person transmission is estimated to be 20% and 80% respectively. Therefore, the proportion of infections environment-to-person transmission *P*_*ie*_ is estimated 0.2.

The detailed parameters for control measures in different intervention strategies were traced using the surveillance data as shown in Table S5.

Finally, a Runge-Kutta algorithm was used to solve the differential equations (1)-(6) to obtain the variations of *S, E*_1_, *E*_2_, *I*_1_, *I*_2_ and *R* with time.

## Data Availability

https://www.who.int/
http://qianxi.baidu.com/

## ACKNOWLEDGEMENTS

The work was supported by the National Key Research and Development Program of China (2016YFC0209000) and the National Natural Science Foundation of China (41375154, 21777193, 41875181, and 41977369). The authors would like to acknowledge Academician Q.C. Zeng for his guidance from the Institute of Atmospheric Physics and Prof. Tunga Salthammer from Fraunhofer WKI for his useful discussion, Prof. J. Li from the Space Science and Engineering Center, University of Wisconsin-Madison, Chinese Academy of Sciences, Prof. Z.F. Yang from the State Key Laboratory of Respiratory Disease, National Clinical Research Center for Respiratory Disease, Guangzhou Institute of Respiratory Health, China, and professors F. Hu, Z.F. Wang, and Z.H. Lin from the Institute of Atmospheric Physics, Chinese Academy of Sciences for their helpful discussions.

## Author contributions

S.H., L.W., and Q.D. designed research; S.H., L.W., L.X., A.Z., L.S., F.L., L.Z., J.L., Y.L., R.H., H.Q., S.F., Z.W., Y.L., T.S., C.L.and Q.D. performed research; S.H., L.W., C.L.,T.S. and Q.D. analyzed data; S.H., L.W., Y.L., and Q.D. wrote the paper.

---

**Table S1.** Values for the parameters in the SEIR-CV model.

| Country | China out of Hubei | The United States | Brazil | India | Five European countries | Northern Hemisphere | Southern Hemisphere |
| --- | --- | --- | --- | --- | --- | --- | --- |
| Total population | 1.34 billion | 0.33 billion | 0.21 billion | 13.24 billion | 0.32 billion | 5.35 billion | 0.83 billion |
| Initial efficiency of quarantine during incubation period, $\lambda_{Q0}$ | 0.000 | 0.657 | 0.364 | 0.582 | 0.306 | 0.293 | 0.523 |
| Initial efficiency of isolation during infection period, $\lambda_{I0}$ | 0.000 | 0.908 | 0.706 | 0.880 | 0.783 | 0.676 | 0.721 |
| Initial efficiency of protection, $\lambda_{P0}$ | 0.000 | 0.347 | 0.198 | 0.482 | 0.106 | 0.247 | 0.278 |
| Initial efficiency of decontamination, $\lambda_{d0}$ | 0.000 | 0.108 | 0.102 | 0.101 | 0.082 | 0.093 | 0.110 |

**Table S2.** Correlation analysis of rolling prediction and surveillance results.

| Name |  | Correlation coefficient | Bias | Average error | Confidence interval(95%) |  |
| --- | --- | --- | --- | --- | --- | --- |
|  |  |  |  |  | Lower limit | Upper limit |
| China out of Hubei Province | Total confirmed cases | 0.99987 | -5.0E-6 | 4.6E-5 | 0.99976 | 0.99993 |
|  | Total confirmed new cases | 0.99180 | 1.7E-5 | 3.4E-3 | 0.98358 | 0.99645 |
| Five European countries | Total confirmed cases | 0.99992 | -2.6E-8 | 1.6E-5 | 0.99989 | 0.99995 |
|  | Total confirmed new cases | 0.95572 | -2.4E-4 | 1.3E-2 | 0.92487 | 0.97598 |
| The United States | Total confirmed cases | 0.99995 | 3.3E-7 | 7.0E-6 | 0.99994 | 0.99996 |
|  | Total confirmed new cases | 0.89934 | -2.8E-5 | 2.7E-2 | 0.83728 | 0.94487 |
| Brazil | Total confirmed cases | 0.99989 | 2.0E-6 | 2.1E-5 | 0.99984 | 0.99993 |
|  | Total confirmed new cases | 0.95064 | 5.0E-4 | 1.1E-2 | 0.92844 | 0.96991 |
| India | Total confirmed cases | 0.99997 | 9.7E-9 | 6.0E-6 | 0.99995 | 0.99998 |
|  | Total confirmed new cases | 0.99658 | 3.0E-6 | 7.7E-4 | 0.99476 | 0.99793 |
| The northern hemisphere out of China | Total confirmed cases | 0.99999 | 4.6E-9 | 2.0E-6 | 0.99998 | 0.99999 |
|  | Total confirmed new cases | 0.98468 | 6.3E-6 | 3.7E-3 | 0.97667 | 0.99110 |
| The southern hemisphere | Total confirmed cases | 0.99991 | 1.0E-6 | 1.6E-5 | 0.99988 | 0.99994 |
|  | Total confirmed new cases | 0.94511 | 1.5E-3 | 1.2E-2 | 0.92025 | 0.96701 |
| The world out of China | Total confirmed cases | 0.99998 | 1.5E-8 | 2.0E-6 | 0.99998 | 0.99999 |
|  | Total confirmed new cases | 0.98510 | 1.8E-5 | 2.7E-3 | 0.97960 | 0.99016 |

**Table S3.** The parameters and physical meanings in the Model

| Parameters | The physical meaning | Value range |
| --- | --- | --- |
| $k_0$ | basic infection rate | $>0$ |
| $k_1$ | the seasonal coefficient of infection rate | $>0$ |
| $\lambda_{Qa}$ | target isolation rate for the persons in the incubation period | $0\sim 1$ |
| $\lambda_{Ia}$ | target isolation rate for the persons in the diseased period | $0\sim 1$ |
| $\lambda_{Q0}$ | initial isolation rate for the persons in the incubation period | $0\sim 1$ |
| $\lambda_{I0}$ | initial isolation rate for the persons in the diseased period | $0\sim 1$ |
| $\mu_Q$ | isolation efficiency factor for the persons in the incubation period | $0\sim 1$ |
| $\mu_I$ | isolation efficiency factor for the persons in the diseased period | $0\sim 1$ |
| $\lambda_{Da}$ | the proportion of the target decontamination | $0\sim P_{ie}$ |
| $\lambda_{D0}$ | the proportion of the decontamination at the initial time | $0\sim P_{ie}$ |
| $\mu_D$ | the efficiency factor of the decontamination | $0\sim 1$ |
| $\lambda_{Pa}$ | the proportion of the target protection | $0\sim 1$ |
| $\lambda_{P0}$ | the proportion of the protection at the initial time | $0\sim 1$ |
| $\mu_P$ | the efficiency factor of the protection | $0\sim 1$ |

$P_{ie}$ is the proportion of infections through the environment

**Table S4.** The course of the disease in the SEIR-CV model.

| The course of the disease | Value |
| --- | --- |
| Non-infectious incubation period, $D_{E1}$ | 2.9 days |
| Infectious incubation period, $D_{E2}$ | 2.3 days |
| Infectious cure period, $D_{I1}$ | 7.0 day |
| Non-infectious cure period, $D_{I2}$ | 14.0 days |

## Supplemental Materials 1

**Parameter inversion method for the infectious disease model with control variables (SEIR-CV)**

The governing differential equations of SEIR-CV are:

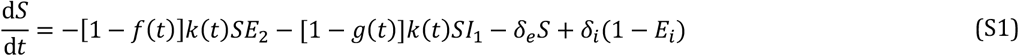

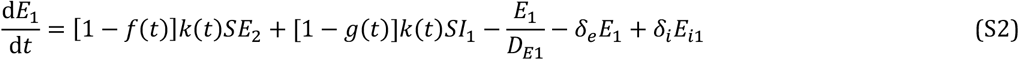

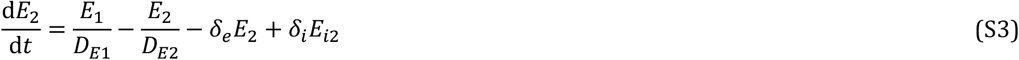

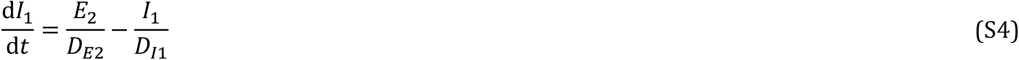

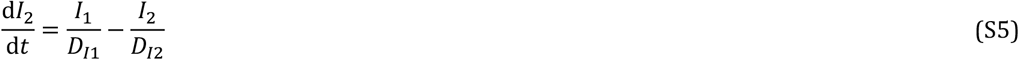

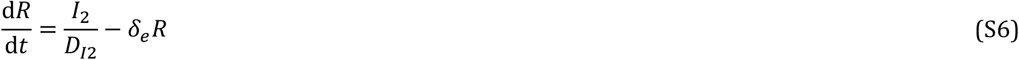

where, *δ*_*e*_ and *δ*_*i*_ are the proportion of emigration and immigration population, respectively; *E*_*i*_ is the proportion of asymptomatic infection from immigration, *E*_*i*1_ and *E*_*i*2_ is the proportion of non-infectious and infectiousness asymptomatic infections respectively. The infection rate,*k*, is considered to vary according to the season, which iis higher during the cold season but lower during the warm season (Figure 1b) ^14-15,27^

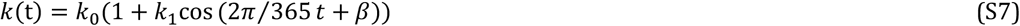

where *k*_0_ is the basic infection rate, related to factors such as the characteristics of the virus and the density of the population, assuming the same value of the asymptomatic and the symptomatic considering there being no obvious difference in viral loads across sex, age groups and disease severity, ^10^ *k*_1_ is the seasonal coefficient.

Contagious function, and *β* is 0 and *π* in the northern and southern hemispheres, respect.ively

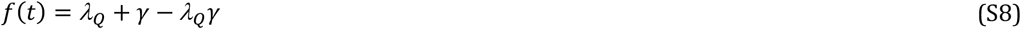

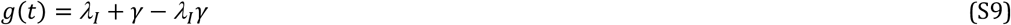

where *λ*_*Q*_, *λ*_*I*_, *λ*_*P*_, *λ*_*D*_ are the efficiencies of the control measures. They are independent of each other, described as follows, Figure 1c:

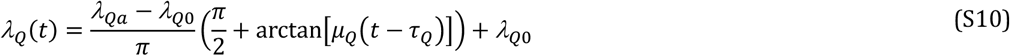

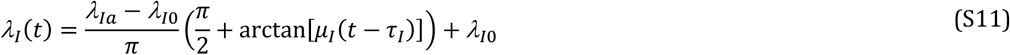

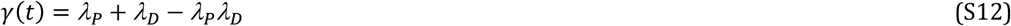

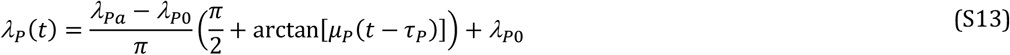

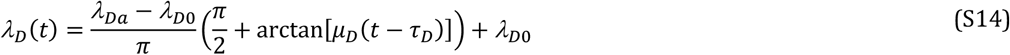

Most of the parameters in the SEIR-CV model cannot be directly determined, they may to be inferred through outbreak surveillance data. In this study, the adjoint method is used for this purpose. Compared with the Bayesian inference, the adjoint method is more efficient and robust with respect to practical usage. The objective function of the adjoint method is established as follows:

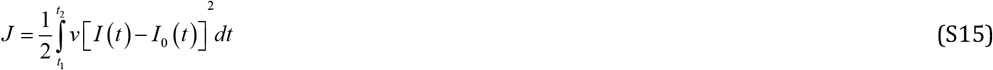

In equation (S15), (*t*_1_, *t*_2_) is the period during which the epidemic develops; *v* is a weighting factor; *I* (*t*)is the proportion of simulated affected cases; *I*_0_ (*t*) is the proportion of statistically affected cases. This objective function can represent the difference between the simulated value and the surveillance data.

The optimal parameters under the constraint conditions should minimize the value of the objective function. This constraint condition is an equation of state, expressed as follows:

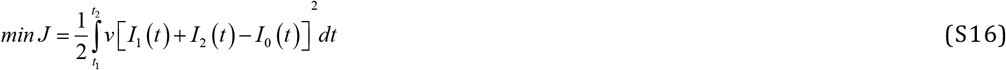

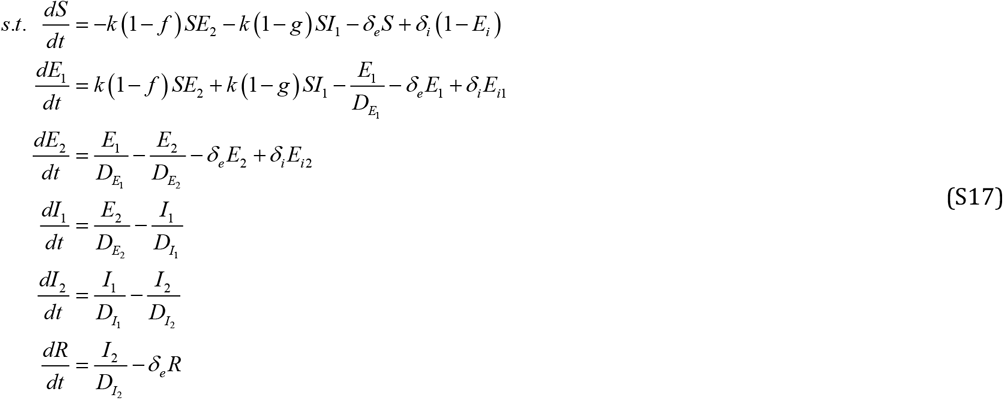

where. *γ*(*t*) = *λ*_*P*_+ *λ*_*D*_ − *λ*_*P*_*λ*_*D*_

To achieve a fast solution, it is necessary to obtain the gradient of the objective function *J* with respect to the model parameters (this gradient is the vector obtained by arranging the derivatives). Iteratively using this gradient information can approximate the optimal solution.

To this end, a Lagrange function is constructed:

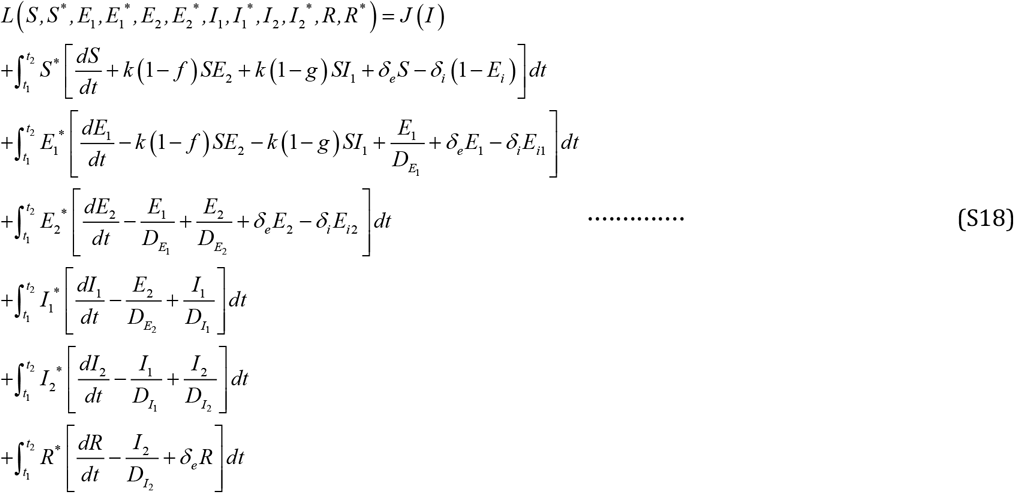

The upper and lower limits of the integral (*t*_1_, *t*_2_) are the periods during which the epidemic develops, and *S* ^*^, *E*_1_ ^*^, *E*_2 *_, *I*_1_ ^*^, *I*_2 *_ and *R*^*^ are the adjoint variables of *S, E*_1_, *E*_2_, *I*_1_, *I*_2_ and *R* respectively. Above these variables are all considered to be independent.

According to the conditions of the above requirements, *S, E*_1_, *E*_2_, *I*_1_, *I*_2_ and *R* minimize the locality of the objective function *J* and *S* ^*^, *E*_1_ ^*^, *E*_2 *_, *I*_1_ ^*^, *I*_2 *_ and *R*^*^ are the necessary conditions for the corresponding adjoint variables (*S* ^*^, *E*_1_ ^*^, *E*_2 *_, *I*_1_ ^*^, *I*_2 *_, *R*^*^, *S, E*_1_, *E*_2_, *I*_1_, *I*_2_, *R*) to be a saddle point of L. So, it must meet the following condition:

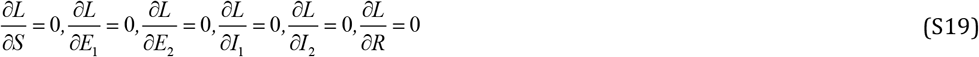

From this, the equation satisfied by the adjoint variables can be derived:

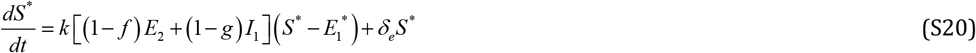

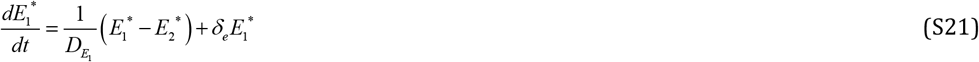

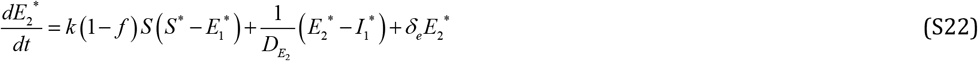

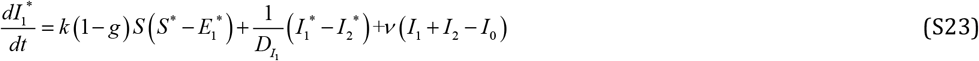

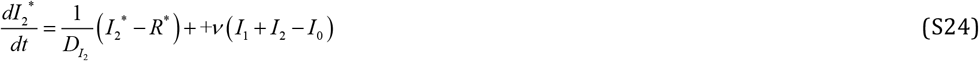

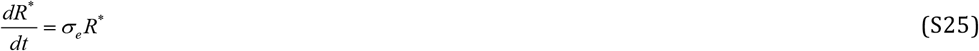

In the case where both the state equations and the adjoint equations are satisfied at the same time, the derivative of the objective function *J* with respect to the model parameter is equal to the derivative of the Lagrange function with respect to the model parameters.

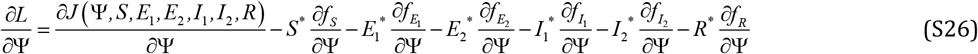

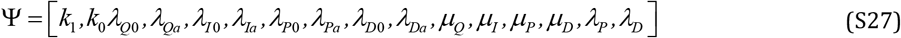

Through the above derivation, the gradient vector *g* can be obtained as:

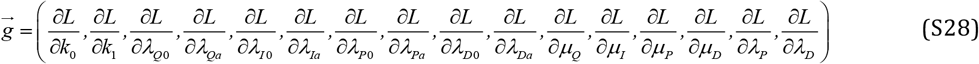

Find the partial derivative for each parameter. The expression for each element in the vector is as follows:

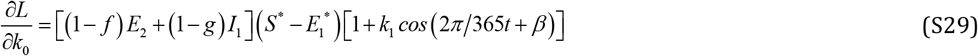

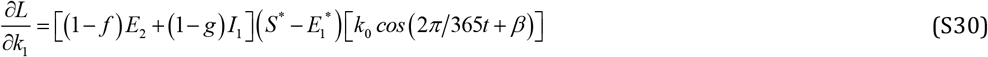

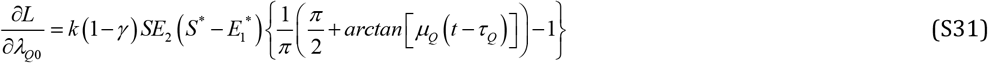

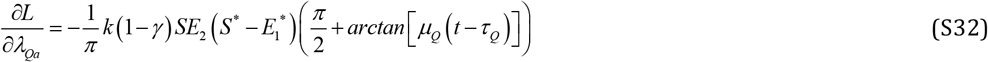

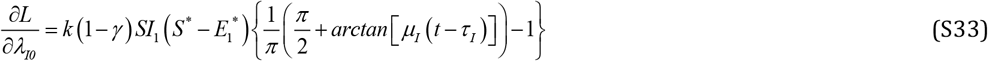

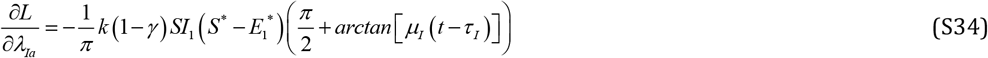

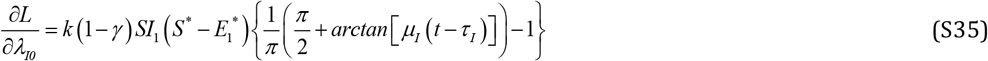

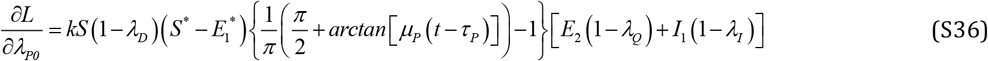

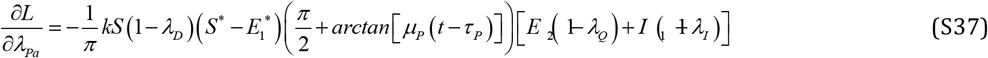

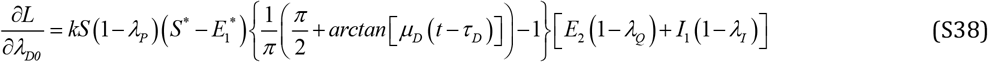

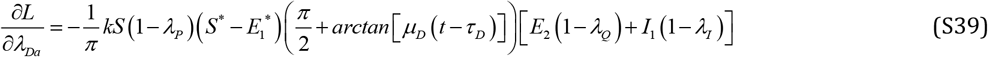

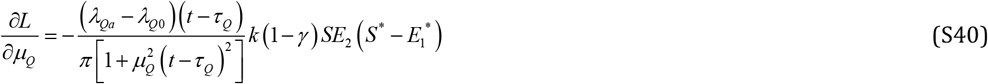

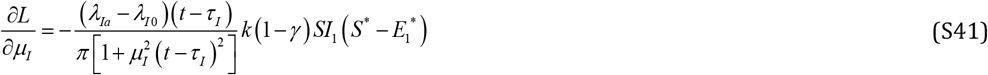

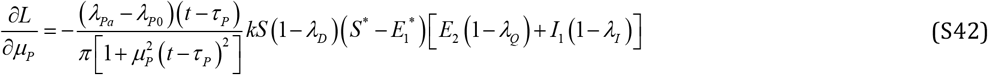

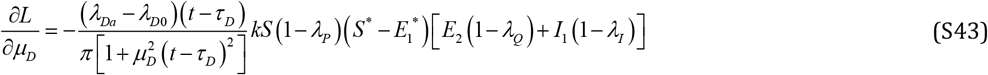

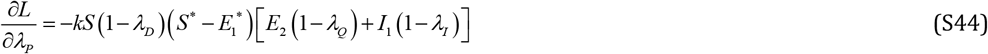

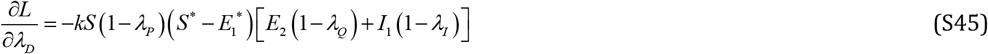

The parameter inversion method based on the adjoint operator method can be summarized as follows:

1. First set initial values for the parameters that need to be inverted;
2. Solving the state equations;
3. Solve the adjoint equations;
4. Find the gradient of the objective function to the model parameters;
5. Determine new model parameter values based on the gradient;
6. Repeat the above steps until the preset conditions for terminating the calculation are reached.

